# Effects of obesity on serum levels of SARS-CoV-2-specific antibodies in COVID-19 patients

**DOI:** 10.1101/2020.12.18.20248483

**Authors:** Daniela Frasca, Lisa Reidy, Carolyn Cray, Alain Diaz, Maria Romero, Kristin Kahl, Bonnie B. Blomberg

**Affiliations:** Department of Microbiology and Immunology, University of Miami Miller School of Medicine, Miami, FL; Sylvester Comprehensive Cancer Center, University of Miami Miller School of Medicine, Miami, FL; Department of Pathology & Laboratory Medicine, University of Miami Miller School of Medicine, Miami, FL

**Keywords:** Obesity, inflammation, antibody responses

## Abstract

SARS-CoV-2 (Severe Acute Respiratory Syndrome Corona Virus-2), cause of COVID-19 (Coronavirus Disease of 2019), represents a significant risk to people living with pre-existing conditions associated with exacerbated inflammatory responses and consequent dysfunctional immunity. In this paper, we have evaluated the effects of obesity, a condition associated with chronic systemic inflammation, on the secretion of SARS-CoV-2-specific IgG antibodies in the blood of COVID-19 patients. Results have shown that SARS-CoV-2 IgG antibodies are negatively associated with Body Mass Index (BMI) in COVID-19 obese patients, as expected based on the known effects of obesity on humoral immunity. Antibodies in COVID-19 obese patients are also negatively associated with serum levels of pro-inflammatory and metabolic markers of inflammaging and pulmonary inflammation, such as SAA (serum amyloid A protein), CRP (C-reactive protein) and ferritin, but positively associated with NEFA (nonesterified fatty acids). These results altogether could help to identify an inflammatory signature with strong predictive value for immune dysfunction that could be targeted to improve humoral immunity in individuals with obesity as well as with other chronic inflammatory conditions.

## Introduction

SARS-CoV-2 (Severe Acute Respiratory Syndrome Corona Virus-2), the cause of COVID-19 (Coronavirus Disease of 2019), has been efficiently spreading from human-to-human since the last months of 2019 and has been responsible for mild-to-severe respiratory tract infections. Our knowledge of human immune responses to SARS-CoV-2 infection is limited, and the host factors responsible for disease progression and symptom severity are largely unknown. Recently published data have indicated that chronic low-grade systemic inflammation, inflammaging [1], is the major cause of the cellular and molecular changes induced by SARS-CoV-2 and is responsible for the highest mortality rates [2]. Inflammaging has been shown to induce chronic immune activation (IA) associated with impairment of immune cell function, as reviewed in [3].

Viral clearance and resolution of SARS-CoV-2 infection requires a complex immune response initiated by resident epithelial cells and innate immune cells, followed by adaptive immune cells that effectively cooperate to eliminate the virus. B cells contribute to viral clearance by producing virus-specific antibodies that can neutralize the virus, thus preventing the spread of infectious virions, controlling virus dissemination, and reducing tissue damage. Previously published data have demonstrated strong neutralizing antibody responses generated against the Spike glycoprotein of the SARS-CoV of the 2002-2003 pandemic protected infected hosts from severe disease [4]. Moreover, it has been postulated that during the current pandemic, the production of antibodies to SARS-CoV-2 is critical to limit disease progression and neutralizing antibodies present in plasma from convalescent COVID-19 patients have been shown to induce fast recovery of critically ill patients [5-7].

Obesity, like viral infections, induces persistent local and systemic inflammation and chronic IA, contributing to functional impairment of immune cells, and decreased immunity. Obesity and associated inflammation lead to several debilitating chronic diseases such as type-2 diabetes, cancer, atherosclerosis, and inflammatory bowel disease [8-15]. Therefore, obesity represents an additional risk factor for COVID-19 patients. Also, the prevalence of disease, as well as the occurrence of complications in obese individuals, are increased as compared to lean controls. Indeed, a strong association has been shown between obesity, obesity-associated comorbidities and severe outcomes of COVID-19 [16]. Retrospective analyses of adult COVID-19 symptomatic patients have demonstrated that individuals with Body Mass Index (BMI) >30 were more likely to be admitted to acute and critical care compared to individuals with a BMI <30 [17]. Previously, it has been demonstrated that obese patients respond poorly to infections [18-20], vaccination [21-23], and therapies [24]. The obesity-associated dysregulation of the immune system may also extend the duration and heighten the magnitude of the metabolic stress. It is well known that the obese adipose tissue (AT) is heavily infiltrated with immune cells [25, 26] which fuel local inflammation and exacerbate inflammaging. AT in the thorax and abdominal areas induce secretion of additional pro-inflammatory mediators that can further compromise lung function [27, 28]. The infiltrating immune cells, once activated following SARS-CoV-2 infection, contribute to the release of inflammatory mediators. Another significant health problem is that the AT may be a viral reservoir, playing a crucial role in maintaining local and systemic inflammation, persistent IA, and immune dysfunction [29].

In this study, we have measured serum levels of SARS-CoV-2 Spike-specific IgG antibodies in lean and obese COVID-19 patients as well as in uninfected controls, using an ELISA test developed and standardized in our laboratory. Results show that higher BMI is associated with a higher infection rate with SARS-CoV-2, measured by serum detection of viral RNA and antibodies. Spike-specific IgG antibodies in obese individuals are negatively associated with BMI and with serum levels of pro-inflammatory and metabolic markers of inflammaging and pulmonary inflammation. Results obtained could help to design an inflammatory signature with a strong predictive value for immune dysfunction that can be targeted to improve humoral immunity in obese infected individuals.

## Results

### Evaluation of Spike-specific IgG antibodies in the sera of study participants using an ELISA standardized in our laboratory

The test group consisted of 52 individuals who were negative, and 72 individuals tested positive for SARS-CoV-2 RNA detection by reverse transcriptase-polymerase chain reaction (RT-PCR) of nasopharyngeal swab samples and serum antibody-positive by lateral flow immunoassay (LFIA) using the lateral flow device (LFD). Age, gender, BMI, routine clinical laboratory measures, and chronic conditions and diseases of the recruited participants are shown in Table 1. Despite several initial reports indicating that COVID-19 patients were characterized by significant lymphocytopenia [30], this cohort had normal numbers of total WBC, neutrophils, and lymphocytes, as also recently shown by other groups [31-33].

**Table 1.**
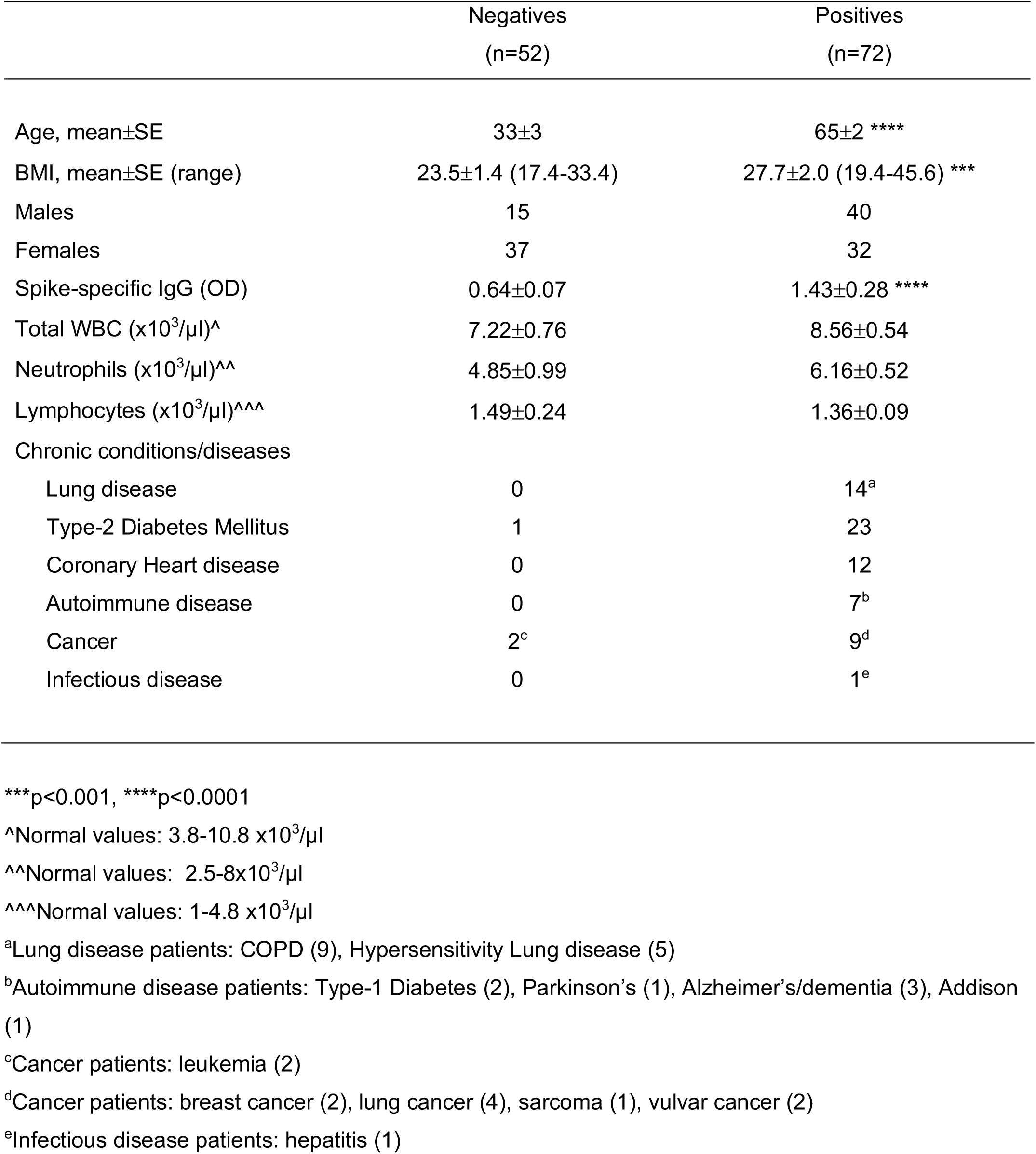
Demographics, laboratory and clinical characteristics of enrolled participants

|  | Negatives<br>(n=52) | Positives<br>(n=72) |
| --- | --- | --- |
| Age, mean±SE | 33±3 | 65±2 **** |
| BMI, mean±SE (range) | 23.5±1.4 (17.4-33.4) | 27.7±2.0 (19.4-45.6) *** |
| Males | 15 | 40 |
| Females | 37 | 32 |
| Spike-specific IgG (OD) | 0.64±0.07 | 1.43±0.28 **** |
| Total WBC (x10<sup>3</sup>/μl)<sup>^</sup> | 7.22±0.76 | 8.56±0.54 |
| Neutrophils (x10<sup>3</sup>/μl)<sup>^^</sup> | 4.85±0.99 | 6.16±0.52 |
| Lymphocytes (x10<sup>3</sup>/μl)<sup>^^^</sup> | 1.49±0.24 | 1.36±0.09 |
| Chronic conditions/diseases |  |  |
| Lung disease | 0 | 14<sup>a</sup> |
| Type-2 Diabetes Mellitus | 1 | 23 |
| Coronary Heart disease | 0 | 12 |
| Autoimmune disease | 0 | 7<sup>b</sup> |
| Cancer | 2<sup>c</sup> | 9<sup>d</sup> |
| Infectious disease | 0 | 1<sup>e</sup> |

\*\*\*p<0.001, \*\*\*\*p<0.0001
<sup>^</sup>Normal values: 3.8-10.8 x10<sup>3</sup>/μl
<sup>^^</sup>Normal values: 2.5-8x10<sup>3</sup>/μl
<sup>^^^</sup>Normal values: 1-4.8 x10<sup>3</sup>/μl
<sup>a</sup>Lung disease patients: COPD (9), Hypersensitivity Lung disease (5)
<sup>b</sup>Autoimmune disease patients: Type-1 Diabetes (2), Parkinson's (1), Alzheimer's/dementia (3), Addison (1)
<sup>c</sup>Cancer patients: leukemia (2)
<sup>d</sup>Cancer patients: breast cancer (2), lung cancer (4), sarcoma (1), vulvar cancer (2)
<sup>e</sup>Infectious disease patients: hepatitis (1)

In addition to previously measured RT-PCR and LFD results, we also measured IgG antibodies specific for the SARS-CoV-2 (2019-nCoV) S1+S2 (Spike) recombinant protein, using an ELISA developed and standardized in our laboratory. This ELISA confirmed the results previously obtained with RT-PCR and LFD as all the individuals tested negative or positive by RT-PCR and LFD were also negative or positive in our Spike-specific ELISA, respectively. Results in Table 1 show significantly higher levels of Spike-specific IgG antibodies in positive *versus* negative individuals in our assay.

### BMI is higher in SARS-CoV-2 positive *versus* negative individuals and is associated with severe respiratory symptoms

Next, we examined the BMI of positive and negative individuals. Results in Fig. 1A show that BMI was higher in positive *versus* negative individuals (27.7 *versus* 23.5, respectively), suggesting a higher frequency of infected patients in obese *versus* lean individuals. In the group of positives, higher BMI was also associated with severe respiratory symptoms (29.1 *versus* 24.6, respectively), as evaluated at the time of hospital admission (Fig. 1B). Severe respiratory symptoms included high fever, cough, shortness of breath, hypoxia, as recorded at the time of hospital admission.

**Figure 1.**
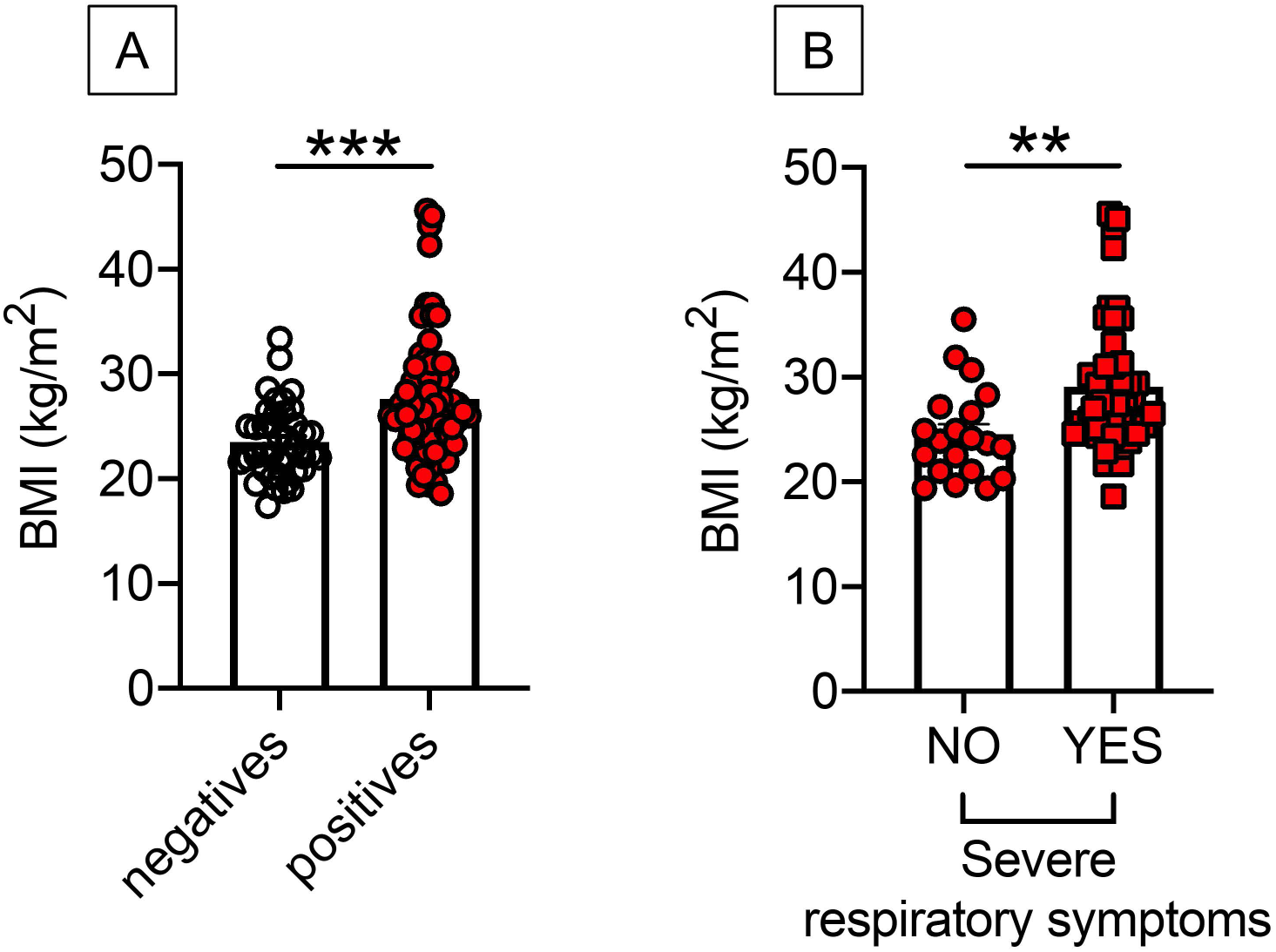
BMI is higher in positive *versus* negative individuals and is associated with severe respiratory symptoms. **A**. BMI in participants tested negative and positive by Spike-specific ELISA. **B**. BMI in COVID-19 positive patients with (n=46) or without (n=20) severe respiratory symptoms. Severe respiratory symptoms included high fever, cough, shortness of breath, hypoxia, as determined at the time of hospital admission. **p<0.01, ***p<0.001.

### SAA, CRP and ferritin, markers of COVID-19, are higher in positive *versus* negative individuals and are associated with severe respiratory symptoms

We measured pro-inflammatory and metabolic markers associated with inflammation in serum samples from positive and negative individuals. We measured SAA (serum amyloid A protein) [34, 35], CRP (C-reactive protein) [36-39] and ferritin [38, 40, 41]. These proteins are markers of pathogen-driven pulmonary inflammation, embolism, and disseminated intravascular coagulation, all characteristics of COVID-19, and therefore predictors of adverse health outcomes. Results in Fig. 2, top show significantly higher levels of these markers in the serum of positive *versus* negative individuals. In the group of positives, SAA and CRP values were also higher in patients with severe respiratory symptoms than those without symptoms, whereas ferritin showed a trend borderline of significance (p=0.06) (Fig. 2, bottom).

**Figure 2.**
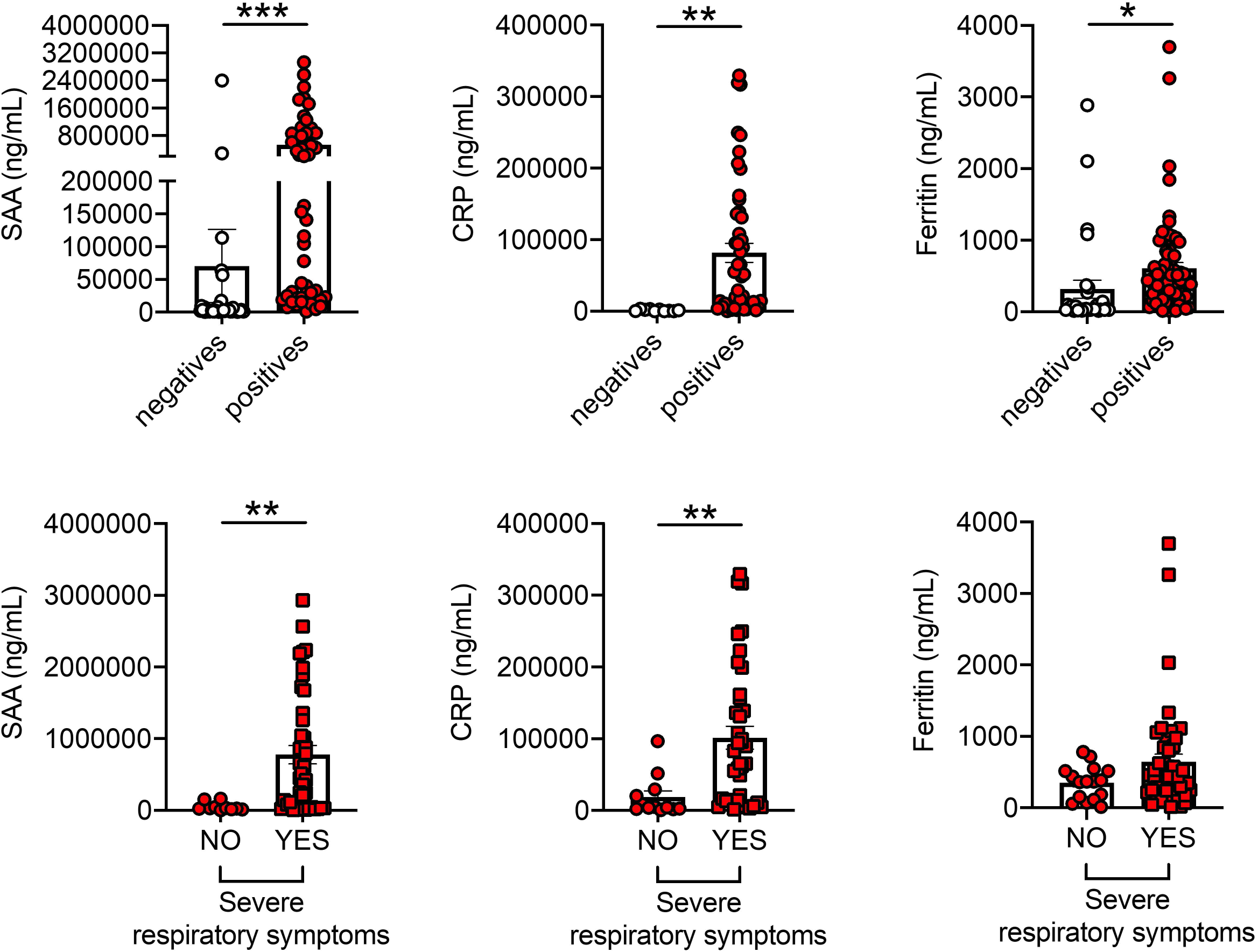
SAA, CRP and ferritin, markers of COVID-19, are higher in positive *versus* negative individuals and are associated with severe respiratory symptoms. **Top**. SAA, CRP and ferritin were detected in serum samples of participants tested negative and positive. **Bottom**. SAA, CRP and ferritin in serum samples of COVID-19 positive patients with [n=42 (SAA), n=38 (CRP), n=48 (ferritin)] or without [n=10 (SAA), n=10 (CRP), n=16 (ferritin)] severe respiratory symptoms (as defined in Fig. 1). *p<0.05, **p<0.01, ***p<0.001.

Conversely, and very interestingly, serum levels of NEFA (nonesterified fatty acids) were found lower in positive *versus* negative individuals and lower in patients with severe respiratory symptoms as compared to those with no symptoms (Fig. 3). These results could be explained by the recently published findings that the receptor binding domain of the Spike protein is physically and tightly bound to an essential fatty acid, the linoleic acid, leading to a locked Spike conformation and reduced ACE2 interaction, at least *in vitro* [42].

**Figure 3.**
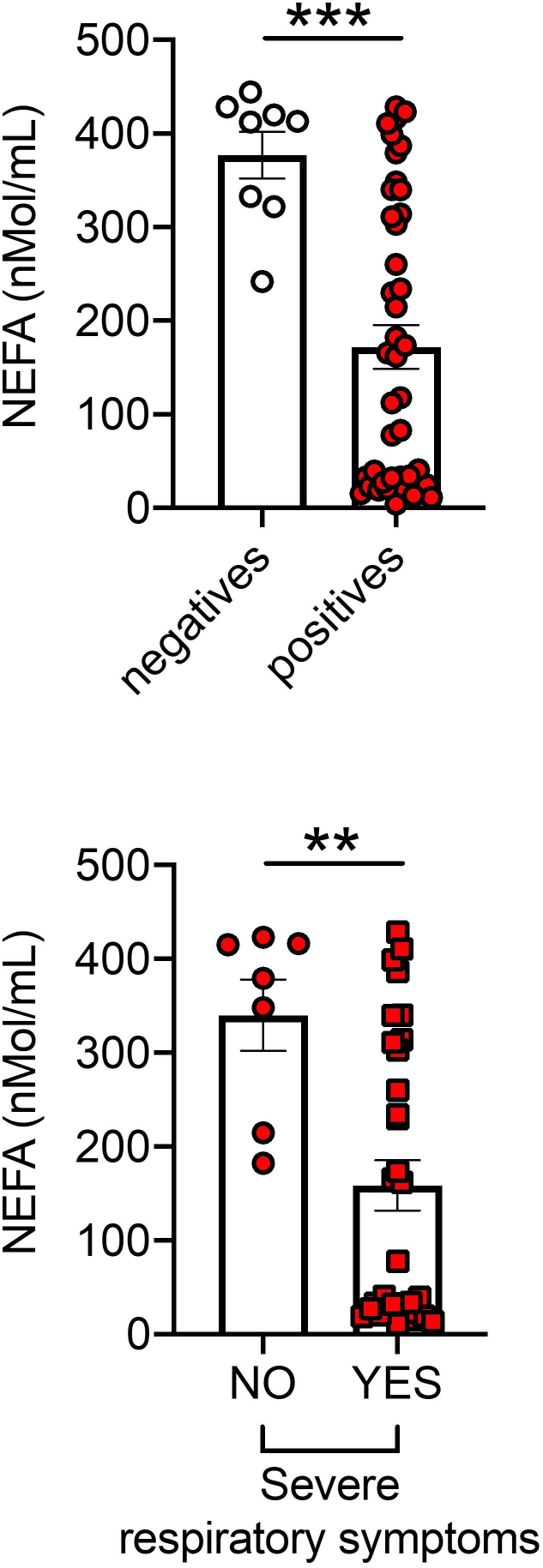
NEFA are lower in positive *versus* negative individuals. **Top**. NEFA in serum samples of participants tested negative and positive. **Bottom**. NEFA in serum samples of COVID-19 positive patients with (n=31) or without (n=7) severe respiratory symptoms (as defined in Fig. 1). **p<0.01, ***p<0.001.

### SAA, CRP, and ferritin are positively associated with BMI and negatively associated with Spike-specific IgG

When we performed correlation analyses between SAA, CRP, ferritin, and NEFA with BMI in positive individuals, we found, as expected, positive associations with SAA, CRP and ferritin, and negative associations with NEFA (Fig. 4). Correlations between serum pro-inflammatory and metabolic markers in the positive individuals are shown in Table 2. Moreover, Spike-specific IgG levels were negatively associated with BMI, SAA, and CRP (Fig. 5, top), and positively associated with NEFA (Fig. 5, bottom).

**Table 2.** Correlations between serum pro-inflammatory and metabolic markers in the positive participants

| Correlation | Person's r | P value |
| --- | --- | --- |
| SAA & ferritin | 0.43 | **0.0029** |
| SAA & CRP | 0.52 | **0.0005** |
| SAA & NEFA | -0.41 | **0.02** |
| Ferritin & CRP | 0.05 | 0.73 |
| Ferritin & NEFA | -0.11 | 0.05 |
| CRP & NEFA | -0.30 | 0.06 |

In bold are indicated the correlations that are significant

**Figure 4.**
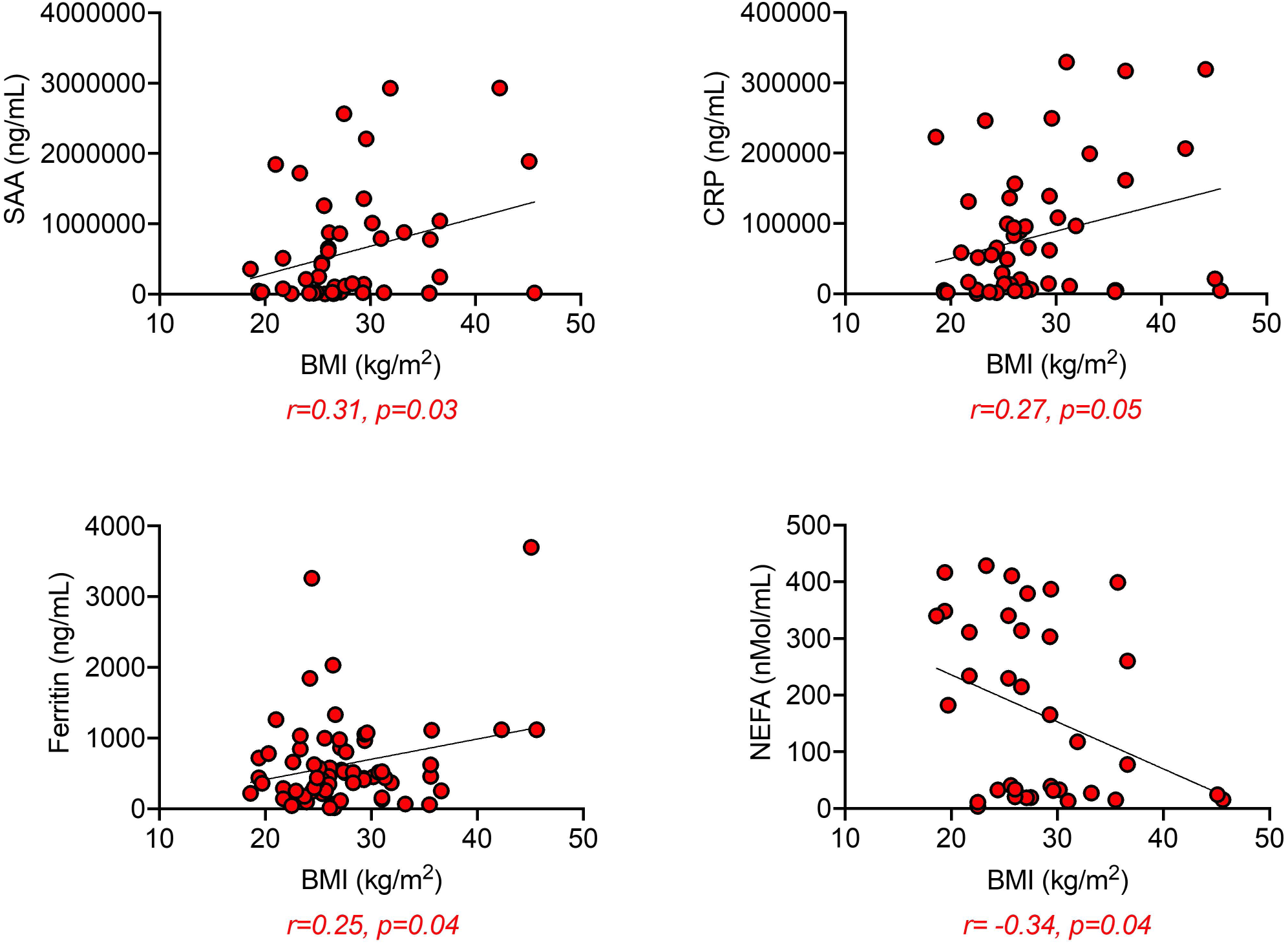
SAA, CRP and ferritin are positively associated, whereas NEFA are negatively associated, with BMI. Correlations of SAA, CRP and ferritin with BMI. Pearson’s regression coefficients and p values are indicated at the bottom of each figure.

**Figure 5.**
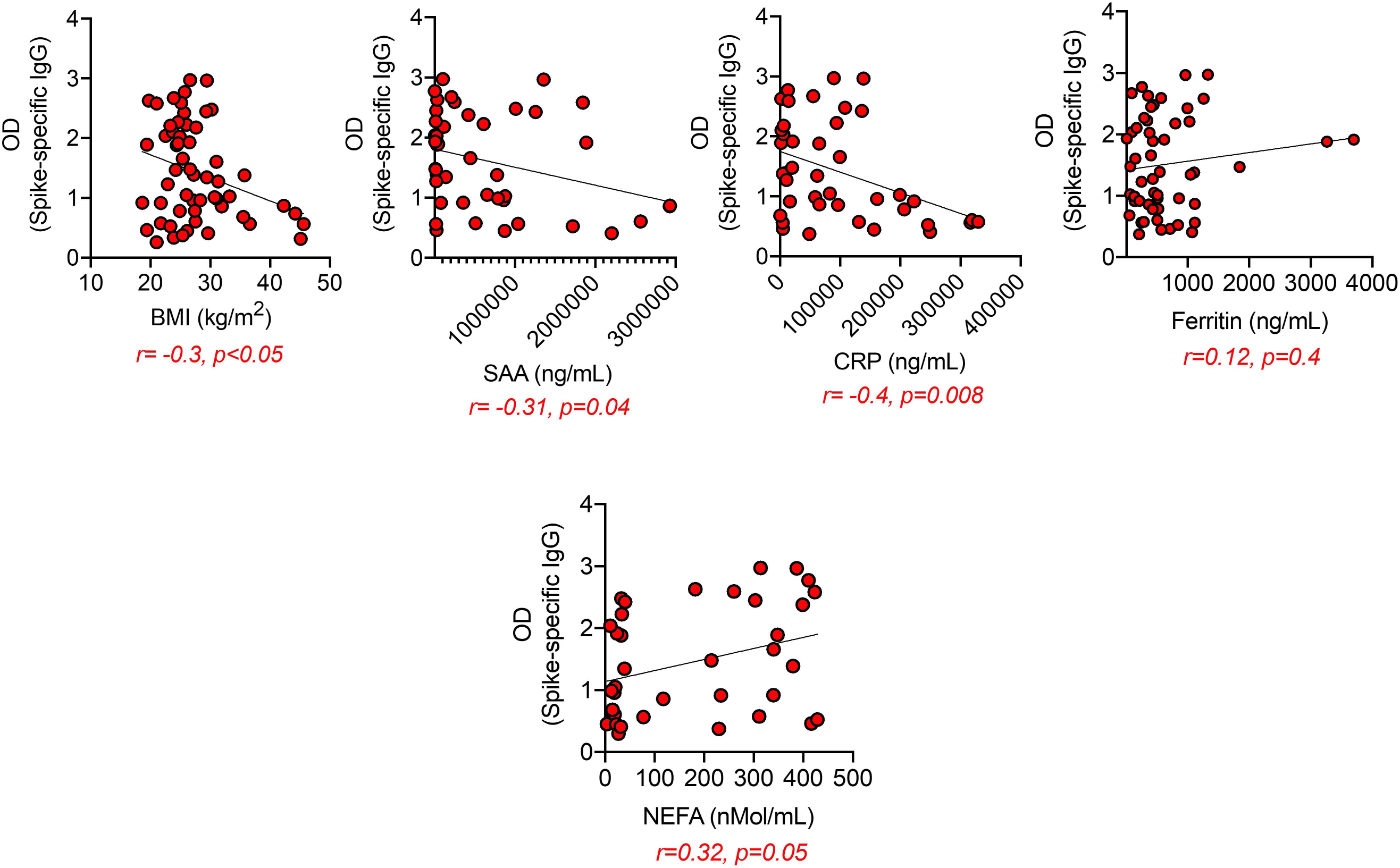
Spike-specific IgG are negatively associated with SAA and CRP and positively associated with NEFA. Correlations of Spike-specific IgG antibodies with BMI, SAA, CRP, ferritin and NEFA. Pearson’s regression coefficients and p values are indicated at the bottom of each figure.

## Discussion

To our knowledge, this is the first study of immunological and inflammatory profiles of COVID-19 obese patients. Although this is a rapidly evolving research field, and it has already been shown that individuals with a confirmed PCR diagnosis of infection develop antibodies against the Spike protein [43], how obesity may affect the secretion of SARS-CoV-2-specific IgG is poorly elucidated. Results herein show that serum levels of SARS-CoV-2 IgG antibodies are negatively associated with BMI in COVID-19 patients. This result is consistent with the knowledge that obesity is an inflammatory condition associated with inflammaging [1] and metaflammation [44] both of which are negatively associated with a functional immune system [45]. Another result from the present study is the negative association of SARS-CoV-2 IgG antibodies with markers of pulmonary inflammation (SAA, CRP, ferritin) in our cohort of COVID-19 patients. These are major inflammatory mediators and markers of inflammatory lung injury in patients with catastrophic acute respiratory distress syndrome, which is a primary consequence of COVID-19. SAA, CRP and ferritin are known to induce a cascade of pro-inflammatory events leading to the secretion of additional markers of inflammation that contribute to the exacerbation of local and systemic inflammation resulting in dysfunctional B cells. In particular, SAA has been shown to induce secretion of several pro-inflammatory mediators by macrophages, such as IL-1β, TNF-α, IL-1RA, IL-8 [46, 47], and IL-33 [48], through activation of NF-kB and IRF7. Similar to SAA, CRP also induces secretion of pro-inflammatory cytokines (IL-6, IL-1β, TNF-α) [49] and chemokines (CCL2, CCL3, CCL4) [50] by monocytes and macrophages, whereas ferritin induces IL-1β and IL-12p70 in macrophages [51]. No direct effects of SAA, CRP and ferritin on B cells have been reported.

Our previously published work has shown that obesity decreases the serum antibody response to the influenza vaccine in young and elderly individuals [21] and increases the secretion of autoimmune antibodies [25, 52, 53]. We have shown that the serum concentration of leptin, the hormone secreted by the AT [54], correlates with the amount of body fat and BMI [55]. This may be at least one molecular mechanism involved in the reduced B cell function in individuals with obesity, as it induces the secretion of pro-inflammatory cytokines (IL-6/TNF-α) in human peripheral blood B cells through activation of JAK2/STAT3 and p38MAPK/ERK1/2 signaling pathways [56, 57]. Leptin also induces intrinsic B cell inflammation as measured by mRNA expression of several markers associated with immunosenescence [58]. Importantly, the expression of these markers in B cells before in vivo/in vitro stimulation negatively correlates with the same B cells’ response after stimulation [59].

We have previously shown that obesity also increases blood frequencies of the subset of double negative (DN) B cells (CD19+CD27-IgD-) [21, 53] which represents the most inflammatory B cell subset. Frequencies of DN B cells also increase in the blood of individuals with inflammatory conditions and diseases. These include aging [60-62], autoimmune diseases such as Systemic Lupus Erythematosus [63-65], Rheumatoid Arthritis [66], Sjogren’s disease [67], Multiple Sclerosis [68], Alzheimer’s disease [69], and pemphigus [70]. An increase of DN B cells have also been reported in the blood of COVID-19 patients and associated with anti-viral antibody responses and poor clinical outcomes, as recently shown [71]. DN B cells secrete pro-inflammatory mediators that contribute to dysfunctional humoral responses and pathogenic autoantibodies that, instead of targeting disease-causing viruses, target infected individuals’ tissues. Anti-phospholipids, anti-type-I interferons, anti-nuclear antibodies and Rheumatoid Factor have been found in a large percentage of COVID-19 patients and linked to severe disease as they may inactivate critical components of the anti-viral response [72]. It is also likely, and important to demonstrate, that the neutralizing antibodies in COVID-19 patients may carry autoimmune specificity, as recently shown for broadly neutralizing antibodies against conserved domains in the influenza hemagglutinin with autoreactivity against tissue antigens previously not identified as autoantigens [73]. Overall, these findings support that the SARS-CoV-2 infection, similar to influenza, may induce self-tolerance breakdown to a variety of autoantigens with self-tolerance breakdown already occurring in obese individuals. It is also likely that tissue failure following dissemination of the virus through the blood can induce cell death and release of intracellular antigens not known as autoantigens, in addition to those already released in the AT for mechanisms like hypoxia and consequent cell death, as we have previously demonstrated [25]. The quality of the antibody response in COVID-19 patients with obesity is extremely relevant for future vaccination campaigns to prevent SARS-CoV-2 infection and COVID-19-associated complications in this population that is likely to be among the first to benefit from vaccination.

## Limitations of this study

There are at least two limitations in our study. First, the number of individuals recruited is limited. We are planning to expand this cohort in the future and with additional markers. Second, this study, although straightforward only shows associations and not mechanisms for the associations. Our future studies will address possibilities after in depth approaches.

## Materials and methods

### Enrolled participants

Experiments were performed using serum samples isolated from individuals tested negative or positive for SARS-CoV-2 RNA by RT-PCR of nasopharyngeal swab samples and by LFIA using the Healgen LFD with specificity for the SARS-CoV-2 Spike antigen. In total, 52 negative and 72 positive serum samples were collected from both inpatient and outpatient settings and frozen until testing was performed. No additional samples were collected from patients, and samples used in this study were only taken for routine clinical care purposes. Samples were de-identified before use in this study. The demographics, BMI, clinical test results, and clinical characteristics of the participants, as well as their SARS-CoV-2-specific IgG ELISA results, are shown in Table 1. This research was approved and reviewed by the Institutional Review Board (IRB, protocols #20200504) at the University of Miami, which reviews all human research conducted under the auspices of the University of Miami.

Both SARS-CoV-2 RT-PCR and serology tests were performed at the clinical laboratories of the Department of Pathology & Laboratory Medicine. SARS-CoV-2 RT-PCR was performed at the University of Miami Hospital using either the Diasorin or the BDMax assay and reagents. Depending on submission type (symptomatic or asymptomatic) the sample was assigned and tested per manufacturer guidelines.

The Healgen LFD serology test was performed according to the manufacturer guidelines. To each cassette 5 µL of serum was added, and two drops of the manufacturer provided buffer solution were applied to the cassette immediately after sample was loaded. Results were read 10 min after. The Healgen LFD were single-channel flow devices, with IgG, IgM, and C (control line) annotated on the cassettes. A pink line in either/or both IgG and IgM was recorded. Because the LFD results are not quantitative, only showing the presence or absence of specific IgM/IgG antibodies, we developed a Spike-specific ELISA (see below) to compare anti-Spike values with the other measures.

### ELISA to measure Spike-specific IgG antibodies

Serum IgG antibodies against SARS-CoV-2 Spike protein were measured by an ELISA developed and standardized in our laboratory. Briefly, 96-well microplates (Immulon 4HBX, Thermo Scientific) were coated with recombinant NCP-CoV (2019-nCoV) Spike protein (S1+S2 ECD) from Sino Biological (#40589-V08B1) at 2 µg/mL for 1 hr at room temperature. Plates were then washed with Tween-20 0.05% in PBS (PBST) and blocked with assay buffer (1% BSA in PBS) for 1 hr at 37°C. All subsequent steps after blocking were performed by a DYNEX DS2® Automated ELISA system (DYNEX Technologies). Serum samples diluted 1:50,000 in assay buffer were added in duplicate and plates were incubated for 2 hrs. Plates were washed with PBST and 100 µL per well of a peroxidase-conjugated goat anti-human IgG (Jackson ImmunoResearch #109-036-098), diluted 1:10,000 in assay buffer, were added. After 1 hr incubation, plates were washed and a stabilized 3,3′,5,5′-Tetramethylbenzidine (TMB) substrate (Sigma) was added to the wells. The enzymatic reaction was stopped after 20 min with a Stop solution (1 M sulfuric acid), and absorbance at 450 nm was read by the DYNEX DS2 instrument.

### ELISA to measure serum pro-inflammatory and metabolic markers

Serum levels of SAA, CRP, ferritin, and NEFA were measured using the following commercially available kits. SAA: Life Diagnostic #SAA-20. CRP: R&D # DCRP00. Ferritin: Thermo Scientific #EHFTL. NEFA: abcam #ab65341.

### Statistical analyses

To examine differences between groups, unpaired Student’s t tests (two-tailed) were used. To examine relationships between variables, bivariate Pearson’s correlation analyses were performed, using GraphPad Prism version 8 software, which was used to construct all graphs.

## Data Availability

The data presented in this study are available upon request to the corresponding author.

## Acknowledgements

This study was supported by NIH awards AG32576 (DF/BBB), AG059719 (DF), and by the University of Miami Department of Pathology & Laboratory Medicine.

## Author Contributions

**Conceptualization:** DF, BBB, LR, CC

**Data curation:** DF, AD, MR, BBB, LR, CC, KK

**Formal analysis:** DF, BBB, AD, MR

**Funding acquisition:** DF, BBB, CC

**Investigation:** DF, BBB, LR, CC

**Methodology:** AD, MR, KK

**Project administration:** DF, BBB, LR, CC

**Resources:** DF, BBB, LR, CC

**Supervision:** DF, BBB, LR, CC

**Validation:** AD, MR, KK

**Writing - original draft:** DF

**Writing - review & editing:** DF, AD, MR, BBB, LR, CC, KK

